# Crossover of the dimensions of work-family and family-work conflict in couples: Protocol for a qualitative study

**DOI:** 10.1101/2023.08.06.23293724

**Authors:** Ewelina Smoktunowicz, Magdalena Lesnierowska, Justyna Ziolkowska, Marta Roczniewska

**Affiliations:** StresLab Research Centre, Institute of Psychology, SWPS University, Warsaw, Poland; Institute of Psychology, SWPS University, Wroclaw, Poland; Institute of Psychology, SWPS University, Sopot, Poland; Procome Research Group, Department of Learning, Informatics, Management, and Ethics, Karolinska Institutet, Stockholm, Sweden

**Keywords:** work-family conflict, family-work conflict, WFC, FWC, crossover, dyads

## Abstract

Conflict between work and non-work is a bidirectional and a multidimensional construct that has garnered much attention from researchers and practitioners alike. Previously, studies with a dyadic design demonstrated that interrole conflict can cross over between partners in romantic relationships. The aim of the present study is to explore—from an individual and dyadic perspective—how partners perceive dimensions of interrole conflict (that is: time, strain, behaviour, and possibly others) and whether crossover between partners is dimension-dependent. This protocol outlines a qualitative interview study. Participants (*N* = 40) will be dual-earner couples that meet two inclusion criteria: both partners need to be professionally active, and the couples need to have lived together for at least a year. Interviews will be conducted separately with each partner. To analyse the data at the individual level we will use reflexive thematic analysis. To analyse the data at the dyadic level we will apply an adapted version of the framework method. We anticipate that findings of this study will have the potential to advance theoretical models depicting crossover processes and, more generally, the interface between work and family lives. Moreover, insights into how couples experience dimension-based interrole conflict will be important for the development of targeted interventions.

## Introduction

Conflict between work and family is a concept that has long interested not only researchers but also, or perhaps mostly, those of us who balance job responsibilities with being a partner, a friend, or a parent. What is less known outside the walls of the scientific community is that interrole conflict does not stop with the person who experiences it but carries tangible consequences for their partner [1]. Greenhaus and Beutell, in their seminal paper [2], defined the work and family interface as “a form of interrole conflict in which the role pressures from the work and family domains are mutually incompatible in some respects” (p. 77). Despite the seemingly straightforward definition, interrole conflict is quite complex. First, it is bidirectional, meaning that we distinguish between work-family conflict (WFC) and family-work conflict (FWC). Second, each of these conflicts is thought to be comprised of three dimensions: time, strain, and behaviour [2,3]. Third, interrole conflict affects not only those who experience it but also the people around them in a so-called crossover process [1]. Hence, for people in relationships, interrole conflict might become a bidirectional and multidimensional dyadic phenomenon.

Distinguishing between six forms of interrole conflict (i.e., time-based WFC and FWC, strain-based WFC and FWC, and behaviour-based WFC and FWC) on the conceptual and measurement level matters as there is evidence that they are differentially associated with antecedents and outcomes [2]. Less is known about the way those conflicts transmit (i.e., cross over) across persons in close relationships, and specifically, whether crossover mechanisms are dimension specific. It is important to know this because capturing mechanisms behind the dyadic transmission of each dimension could advance theoretical models of crossover and allow them to make better predictions on how interrole conflict of one person in a dyad affects another one. Furthermore, identifying those forms of interrole conflict that are the most detrimental for the well-being of others and the way they transmit might help couples create strategies to address the problem. Subsequently, that might improve their outcomes both at home and at work, such as stress and satisfaction with various domains of life. In the current study we apply qualitative methods to explore dimension-based interrole conflict and we pay particularly close attention to the process of its crossover between partners in dual-earner couples.

### Interrole conflict: directions and dimensions

WFC and FWC vary in their relationships with predictors and outcomes. Meta-analytical findings show that while WFC and FWC are associated with both work and family demands, the effect sizes are larger *within* domains, that is, WFC is predominantly related to work, and FWC to the family/non-work context [4,5]. Conversely, while a cross-domain pattern was expected for the consequences of interrole conflicts (with WFC mostly affecting the non-work domain, and FWC the work one), data indicates that the consequences of WFC and FWC span across all domains. Although meta-analyses have found that WFC is actually more closely linked with work outcomes, and FWC with family ones [6–8], ultimately, they predict a variety of outcomes, including those that do not clearly belong to one domain only, such as health. Moreover, although gender differences in both WFC and FWC are considered too small to bear meaningful consequences, a meta-analysis has shown that the effect is slightly larger for FWC than WFC: It is more often reported by women [9]. Finally, a genetic component in interrole conflict explains more variance in WFC than in FWC but, at the same time, controlling for dispositional factors does not significantly reduce the relationship between job demands and WFC, whereas it does for the relationship between family demands and FWC [10]. Study authors suggest that people might be better at looking at job demands objectively and identifying when they interfere with their family life, but their perceptions of family demands and FWC itself overlap. Taken together, these findings require that both directions of interrole conflict be investigated.

WFC and FWC are even more nuanced; they are conceptualised to be comprised of three dimensions: time, strain, and behaviour [2,3]. *Time-based conflict* is probably the most intuitive one and occurs when there are pressures in one role that lead to people being either physically unable or too preoccupied to perform in another one. People experience *strain-based conflict* when demands in one role leave them too anxious or fatigued to meaningfully take part in their other life roles. Finally, a need to behave differently at home and at work, due to external standards, may lead to *behaviour-based conflict*. A model proposed by Greenhaus and Beutell [2] and empirical evidence indicate that these dimensions show varying associations with their predictors and outcomes [11–13]. There also seem to be gender differences [9]. Specifically, men report experiencing more time-based WFC than women, and women report more strain-based WFC and FWC than men. Importantly, interrole conflict dimensions have not received equal attention as there is more data on time and strain dimensions than on behaviour [13]. It is possible that behaviour-based conflict is less intuitive for people to evaluate, and hence it is imperative to find a way to explore it and deepen our understanding of it.

Potentially, there are more dimensions of interrole conflicts. For example, Greenhaus et al. [14] added an *energy* dimension to reflect that performance in one role can be diminished in another due to exhaustion. This would make it different from strain-based conflict, which is a manifestation of symptoms such as irritability, anxiety, and apathy [2]. Kengatharan and Edwards [15] proposed that the three existing dimensions did not cover being too preoccupied (emotionally and cognitively) with one life domain to perform adequately in another. Hence, they proposed adding a *psychological* dimension. Yet, Greenhaus and Beutell [2] previously included role preoccupation in the time dimension. What stems from these propositions is that the dimensional structure of WFC and FWC might need revising [13]. One way to do this is to identify manifestations of interrole conflict and see whether they surpass what is captured by the current dimensions of time, strain, and behaviour.

### Crossover

Twenty years after introducing the original definition of work-family conflict, Greenhaus et al. [14] proposed refining it to highlight that we can only speak about interrole conflict when “(…) experiences in the work (family) role result in diminished performance in the family (work) role” (p. 65). This emphasis on reduced performance seems particularly important when we look at the dynamics of interrole conflict in couples. When one person in a dyad experiences work-family or family-work conflict, this might become salient to the other person only when that experience leads to a less than satisfactory contribution to family life. For example, if a man struggles due to emotionally draining work but still manages to meet demands at home, such as completing household chores or caring tasks, this stress would not be seen as strain-based interrole conflict and might go unnoticed by the man’s significant other and thus not affect them. However, if the same work demands prevent him from performing all these tasks on the family front, then, what we define as strain-based WFC might affect his partner. This across-individuals transmission of experiences, such as stress and interrole conflict^1^, is called crossover [16,17].

In her influential paper, Westman [17] proposed that crossover occurred via one of three routes. The first is a direct empathic crossover: When one partner experiences stress, it initiates an empathic reaction from their partner. The second potential mechanism is called a common stressor: When both parties share a stressor (e.g., a sick child) it is likely to elicit a response from both of them. Finally, crossover can be indirect, that is, an effect of one partner’s stress on another one’s well-being can be mediated and/or moderated by factors such as coping strategies, communication strategies, social support, or social undermining. In their spillover-crossover model (SCM), Bakker and Demerouti [1] propose that negative states and experiences from work can flow to the family/non-work domain via work-family conflict (spillover), leading to loss in well-being. Subsequently, the other person in a dyad becomes affected (crossover) either because negative states cross over directly between partners or indirectly, via mechanisms such as social undermining. Further research on those potential mechanisms in the realm of interrole conflict [18] has highlighted the role of negative marital interactions. Furthermore, meta-analytical findings [19] have confirmed both the direct crossover between a person’s WFC and their partner’s psychological distress and family satisfaction, as well as an indirect one; specifically, people who experience WFC tend to reduce their positive social behaviours which is linked to their partner’s well-being. What is missing in this exchange though, is how partners react to those behaviours in terms of their own thoughts, emotions, and behaviours. Moreover, although common stressors as a crossover mechanism have not been investigated specifically, Li et al. [19] acknowledged that some of their findings could be attributed to this mechanism.

There are some important gaps in the current state of the art. First, FWC has been less researched overall, including the studies on crossover. The spillover-crossover model (SCM) [1] focuses on the flow of experiences from work to family. The authors acknowledge that the transmission can also occur with family demands spilling over to the work domain but when it comes to crossover, they point only to other employees as recipients in this transmission. This is in line with the SCM premise that for the crossover to take place people need to share the domain. Yet, there is some empirical evidence suggesting that FWC might also affect people at home. First, as we mentioned earlier, meta-analytical data show that FWC, just as with WFC, to a certain extent, has consequences in all life domains, including the family domain. Those consequences, in the form of stress or decreased satisfaction with non-work life might affect people at home. More directly, Smoktunowicz and Cieslak [20] found in their dyadic study with heterosexual couples that men’s family-work conflict mediated the relationship between men’s family demands and women’s family-related perceived stress. Yet, the authors did not test for the mechanisms behind this transmission. In sum, although we have some insight into how the crossover of WFC occurs, we know substantially less about how this process looks for FWC. Second, while the dimensions of interrole conflict manifest varying associations with their antecedents and outcomes, no study has explored whether this dimensionality matters in the crossover process. For example, when one partner is often late from work due to external pressure to put in long hours and is hence not present for the family, their partner might be forgiving of this reduced performance, knowing that ‘the fault’ lies elsewhere. Under such circumstances, it might be less likely that this time-based conflict will cross over through negative family interactions, but it might do so through another mechanism, namely empathy. On the contrary, it is plausible to expect that when behaviour-based WFC manifests in underperformance as a spouse or a parent, this will result in tensions, and subsequently family member well-being will deteriorate. With this qualitative study we aim to explore whether, in their stories about work and family, partners perceive these nuances in interrole conflict and react differently to the various forms it takes.

### Study aims

This study aims to explore the subjective experiences of WFC and FWC in a dyadic setting. The primary goal is to investigate whether, and if so, how the dimensions of WFC and FWC cross over between partners in their narratives about their private life and work. Specifically, we formulated two research questions.

RQ1: What dimensions of WFC and FWC do partners identify in a) themselves and b) their partners?

RQ2: How do different dimensions of WFC and FWC cross over between partners?

The secondary goal for this study is to inform a planned internet intervention [21] that will aim to reduce WFC and FWC at the individual and dyadic level. We know that people utilise strategies to cope with interrole conflict [22] including negotiating a division of labour, outsourcing demands and seeking social support. Yet, we are not aware of any evidence-based program that would directly target couples’ WFC and FWC. To inform such a program we aim to identify the strategies that dual-earner couples already employ and the situational factors that can facilitate or prevent their participation in an internet intervention. This goal translates into the following two research questions:

RQ3: What coping strategies with WFC and FCW on an individual and dyadic level do partners identify?

RQ4: What potential barriers and facilitating factors in participating in an internet intervention designed to reduce WFC and FWC do partners recognise?

## Materials and methods

### Study design

This research will employ an experiential qualitative approach to gain an in-depth understanding of lived experience of conflicts between work and family (WFC and FCW) and how they cross over between partners [23]. The decision to use an experiential approach, results from the study goal of gaining a bottom-up perspective on conflicts between work and family and their crossovers to provide a more nuanced understanding of them. The theoretical paradigm underpinning this research is post-positivism [24]. We assume that knowledge has a provisional nature and understanding of reality is always limited and subject to change. Still, through reflexivity and careful methodological procedures, we can aim for a reliable understanding of reality [25].

This protocol follows the Standards for Reporting Qualitative Research [26] and has been preregistered at the Open Science Framework: https://l1nq.com/tLY3E. The study was approved by the departmental Ethical Review Board (decision number 32/2023 issued on July 11^th^, 2023). Participation in this study is voluntary and study participants can withdraw at any time. Informed consent that outlines the purpose of the study, the procedures involved, participants’ rights as well as information about confidentiality, anonymity and the storage of the data, will be obtained from each individual before the commencement of the interview.

### Participants and recruitment

We plan to recruit 20 heterosexual, dual-earner couples. Sampling will be convenient with inclusion criteria ensuring that we will get a sample that is likely to actually experience a conflict between work and life, that is, a sample of couples who are professionally active and live together. Specifically, both partners need to be employed, work a minimum of 50%, and have lived together for at least a year. These criteria are often applied in dyadic studies on interrole conflict [20]. We will recruit couples both with and without children and those that live in various places across the country. We do not set any criteria when it comes to partners’ age, relationship length or organisational tenure, yet couples will be excluded if one of the partners is under the age of 18, refuses to participate in the study or does not speak fluent Polish. To determine the sample size, we used the concept of information power [27,28]. We estimated sample size based on: (1) the specificity of study aims, (2) the specificity of the study sample, (3) the theoretical background of the study, (4) the quality of the interviews with study participants and, finally, (5) analysis strategy (cross case vs. case). To avoid confirmation bias and selecting participants who support our expectations, the recruitment of participants will be performed in collaboration with a specialised research agency, which will facilitate access to a relatively diverse sample in terms of inclusion criteria and allow for participant reimbursement while keeping their data anonymous to the researchers. Recruitment will begin as soon as the study protocol is accepted.

### Data collection

Data will be collected through semi-structured interviews [29] with both members of a couple, but separately. We chose the semi-structured interview because it allows for in-depth subjective perspectives from the interviewees, while providing a consistent framework for all interviews to increase the reliability of the data [30]. The decision to use separate rather than dyadic interviews was based on a literature review of the different types of interviews in dyadic studies [31–35]. As we aim to capture the individual perspective within the dyad without forging a couple perspective, we decided to collect individual interviews, which allow subjective views to be obtained and the opportunity for self-reflection [31,36]. We plan to conduct the interviews with partners simultaneously by two members of the research team, but if this turns out to be unfeasible for a given couple, we will conduct them consecutively, aiming to prevent partners from discussing their interviews beforehand.

The semi-structured interview guide consists of a series of open-ended questions designed to elicit the subjective perspectives of the study participants (see a table in S1). The guide was developed on the basis of a literature review and the study objectives and includes main questions as well as possible follow-up questions. This will allow the interviewer to maintain flexibility during the interview by deepening the interviewee’s narrative and/or developing new topics that emerge during the interview.

Given the characteristics of our study group (working age group), the widespread use of online communication during the Covid pandemic, combined with the advantages of online research interviews (e.g., the possibility of recruiting participants from different parts of the country, cost and time efficiency as well as accessibility), we decided to conduct interviews online using virtual communication platforms and applications such as Skype or Zoom. As we are aware of the challenges associated with such communication channels (e.g., privacy issues, challenges in creating an intimate and private research setting), technological limitations (e.g., access to equipment and software and skills to use them), as well as challenges related to establishing rapport and maintaining contact (for an overview of advantages and disadvantages of online interviewing and recommendations, see, e.g., [37, 38]), we plan to implement a number of precautionary measures to mitigate the potential risks. Firstly, the interviews will be conducted by an experienced researcher who has received training in qualitative interviewing techniques and is competent in establishing rapport (including in online settings), using probes, listening and maintaining confidentiality. Secondly, in terms of technological challenges, study participants will be able to choose their preferred platform or application for data collection. They will also receive written instructions on how to use the platforms/applications. Finally, in terms of providing a safe, private and quiet space for an interview, participants will be given instructions on how to prepare for an interview. For example, they will be instructed on how to set a blurred or artificial background on the platform to ensure privacy. In addition to the interviews, we will ask the participants to fill out a demographic survey with basic questions about gender, age, being a parent, and job type, as well as a scale to measure dimension-based interrole conflict [39]. The objective of the latter is to gather descriptive data about the sample, i.e., to what extent did they experience interrole conflict.

Transcription of the interviews will be anonymised to protect the identity of the study participants, and the audio recordings will be deleted immediately after transcription. Transcriptions will be stored in password protected files that will be accessible only to the research team. Importantly, signed informed consent will be stored separately, in another password protected file, and it will not be possible to link them to the transcripts.

### Data analysis

Analyses will be conducted at the individual and dyadic levels. We will employ two forms of thematic analysis (TA) [25] which is a widely used qualitative method that allows for a nuanced understanding of rich and complex data by identification, analysis, and interpretation of meanings within qualitative data [40]. At the individual level we will be guided by the principles of reflexive TA [40]. As we are interested in partners’ lived experiences of complex and multifaceted conflicts between work and family (WFC and FCW), reflexive TA is best suited to obtaining such a bottom-up individual perspective. We will apply reflexive TA to all research questions. With regard to Research Question 1 (RQ1), we will analyse which dimensions of WFC and FWC (i.e., time, strain, behaviour and possibly others) partners identify in themselves and in their partners, e.g., are there differences in how study participants see WFC and FWC in themselves and how they see them in their partners? In relation to Research Question 2 (RQ2), reflexive TA will help us to explore lived experiences of the transmission of different dimensions of WFC and FWC between partners. We will also use reflexive TA to analyse statements about coping strategies at the individual and dyadic level (RQ3), e.g., do partners differentiate strategies according to the level (individual vs. dyadic) or dimension of FWC and FCW? Finally, reflexive TA will also be used to elicit partners’ perspectives on barriers and facilitators to participating in an internet intervention designed to reduce WFC and FWC.

The analysis will be carried out in six stages, as recommended by Braun and Clarke [40,41]. Phase one will be preceded by a transcription of the data and, as part of this phase, team members will familiarise themselves with the data. The second phase will involve the production of preliminary codes from the data. We will use the MAXQDA software to facilitate the analysis. In phase three, two members of the team (ES as a researcher of work-life conflicts and JZ as a qualitative researcher) will review the codes and look for a pattern of meaning (initial themes) from coded and collated data. In the next phase, a revision of the preliminary themes will be undertaken to ensure their correspondence with the codes and patterns identified in the dataset. The fifth phase involves defining and naming themes. So, following Braun and Clarke [41], an entire research team will seek to identify the ‘essence’ of what each theme is about. Finally, in the writing-up phase, the themes will be linked in a coherent narrative.

At the dyadic level, we will use the framework method (FM) [42,43] as adapted by Collaco et al. [44]. The FM was chosen because we wanted to complement the analyses at the individual level with a perspective within couple dyads. Thus, the use of the framework method will add to the reflective TA and answer the question of whether the identification of WFC and FWC dimensions match across partners’ narratives (RQ1). In relation to RQ2, the framework analysis will allow us to see how different dimensions of WFC and FWC cross over between partners in dyads (RQ2) and what similarities and differences there are in the stories of coping strategies within dyads (RQ3). The latter will be a very important perspective in the context of the planned intervention. A key advantage of FA is that it allows data to be compared within dyads, with the analysis focusing on overlaps and contrasts in the narratives of the couples. In addition, being more structured and systematic than a reflexive thematic analysis, FA is also efficient and easy to apply even for inexperienced qualitative researchers [40]. The FA process is based on the adapted by Collaco et al. [44] version of the framework method [42,43]. The adapted dyadic analysis using the framework method consists of eight stages, the first three of which (transcription, familiarisation with the data and development of a data coding system) are consistent with the steps of reflexive TA. In the next step we will produce tables of themes and sub-themes, including dyadic codes/summaries for each couple. Step 5 analyses of the degree of agreement between the partners in the couple and how each theme influenced the other. On this basis, dyadic codes will be created. Stages 6 and 7 will involve the creation of new themes based on the dyadic sub-codes and then apply them throughout the corpus. The final stage is to analyse and interpret variations in dyadic couples in reference to a theoretical model. As in the reflexive thematic analysis, the leading roles in the analysis will be played by ES and JZ, with the other team members stepping in as a reflexive team.

### Trustworthiness

An important part of our study design and analysis is to ensure the trustworthiness of our research [44]. To enhance the credibility, transferability, dependability, and confirmability of our study, we follow established procedures in qualitative research [44,45]. Above all, it is the process of reflexivity [46]. Our research team is diverse in terms of past interests, preconceptions, standard methodology and personal experience. This translates into discussions about the differences between us and their potential impact on the study design, data collection and analytical process. In order to maintain the process of reflection, we will meet regularly to discuss differences between us in relation to the research process. In addition, we plan to incorporate a wide range of strategies to ensure the trustworthiness of our research, as proposed by Nowell and co-authors [44]. For example, we plan to involve all team members in key stages of the research process. We plan to collect and provide readers with rich, detailed and contextual information about the dataset and the subsequent stages of the research process. The research protocol serves as a peer debriefing. In the later stages of the research, we also plan to use peer debriefing [45] to get an external perspective.

## Discussion

This paper presents a protocol for a qualitative study aimed at reporting on couples’ experiences with conflict between work and family. Specifically, we are interested in whether people in relationships identify time, strain, behaviour and possibly other dimensions in their own and their partners’ experiences of interrole conflict, and in how those dimensions cross over within couples. Identification of the crossover mechanisms behind dimension-based WFC and FWC is important for the advancement of theoretical models: Those that pertain directly to crossover, such as the spillover-crossover model [1] and those that attempt to depict work and family lives more broadly, such as the job demands-resources model [47]. For example, if FWC dimensions of one person in the relationship carry consequences for their partner—directly and/or indirectly—it might imply that SCM should be extended with yet another dyadic exchange. Furthermore, if one of the WFC or FWC dimensions turned out to be more salient than others to partners in a dyadic setting, it would suggest that demands behind such a dimension have far reaching consequences; knowing this would be important from a theoretical but also a practical standpoint. Traditionally, research on mitigating interrole conflict has focused on the organizational side, in particular on flexible work [48]. While investigating these solutions is essential and should be continued, it is imperative to simultaneously look for means to manage interrole conflict on the family front. This could be achieved through programs dedicated to supporting working couples, and one way to deliver those is online. Internet interventions [49–51] have gained significant popularity over the past three decades with randomised controlled trials repeatedly demonstrating that they are effective in improving people’s well-being, including in the context of work [52,53]. Yet, to the best of our knowledge, no such intervention has targeted interrole conflict and its crossover directly. Findings from this qualitative study have the potential to inform the content of the ensuing intervention, in particular, when it comes to providing examples of how the conflict tends to manifest itself in order to help participants identify the signs and support their partners. Internet interventions are highly accessible, ensure privacy, and can be scalable, which is critical considering that access to evidence-based psychological support is limited [54]. On the downside, depending on their design, these interventions can suffer from low adherence and high dropout [55]. It is crucial to foresee and address barriers that potential participants might face, and to enhance those conditions that make it easier for them to join. Hence, our secondary goal in this study is to collect information on what can persuade couples to participate in interventions that are delivered online and dedicated to coping with interrole conflict.

This study has a few potential limitations. First, our study aims require that we explore interrole conflict at the granular level. However, differentiating between directions of the conflict (i.e., WFC vs. FWC) and particularly its dimensions (i.e., time, strain, behaviour and possibly others) might prove difficult for the interviewees. We plan to address this in two ways. Couples will fill out an interrole conflict scale before the interview which will familiarise them with this concept and provide time to reflect. Additionally, we will conduct pilot interviews to test the interview guide to ensure that our questions are clear and that we are able to meet study goals. If necessary, we will modify the interview scenario. The second limitation is that we will only focus on the negative aspects of balancing work and family life, whereas crossover of enrichment is also possible [56]. We choose to limit our investigation due to feasibility but acknowledge that capturing the work-family interface in its full complexity requires taking into account not only the losses but also the gains. Possible limitations are also related to the design of the study and qualitative methods of data analysis. The main challenge concerns recruitment. Recruiting couples where both partners are willing to share their experiences can be challenging [33,57], particularly as the research interview may involve difficult experiences and couples may be reluctant to share details of work-family conflicts in their relationship. To mitigate the risks of bias in the selection of couples for the study due to potential unwillingness to participate in the study, recruitment was delegated to an external agency. Recruitment by an external agency will also allow respondents to feel more anonymous, as the researchers will not have access to their personal information. Furthermore, given that research interviews may involve difficult experiences for the couple being interviewed, there are challenges associated with the interview and the process of managing confidentiality between partners [58]. Although we will encourage study participants to maintain their privacy during the interview, we cannot rule out the possibility that participants may overhear each other if there are no conditions for simultaneous interviews, or that they may not consult with each other to establish a shared narrative of their experiences. It will therefore be crucial to ensure that, prior to the interviews, participants receive detailed information about the aims and ethical principles of the study, including how we will manage confidentiality. Equally important will be the competence of the experienced interviewer to manage confidentiality, especially in the face of competing accounts in subsequent interviews with partners. Finally, as we are using two methods to analyse the data at two levels - individual and dyadic - this presents challenges for the analytical process with a potential risk of oversimplifying complex phenomena and reducing the richness of the data [40]. In order to avoid errors within the research process, a number of trustworthiness strategies are planned.

## Conclusion

In sum, this study has the potential to provide a more nuanced understanding of the interface between work and family in dual-earner couples. By performing qualitative interviews with couples, we will gain a more-in-depth knowledge on the facets of the conflicts between the work and non-work domains, as well as how they transfer (cross over) between partners. As a consequence, not only theoretical models could be refined and extended, but also practical implications about how to mitigate the consequences of conflicts and their crossover could be developed for future interventions.

## Data Availability

No datasets were generated or analysed during the current study. All relevant data from this study will be made available upon study completion.

https://l1nq.com/tLY3E

## Footnotes

1 Although outside of the scope of this study, we want to highlight that a crossover of positive experiences also woccurs [e.g., 18, 1].

